# *DIP2B* CGG repeat expansion in siblings with neurodevelopmental disability and progressive movement disorder

**DOI:** 10.1101/2024.06.05.24308127

**Authors:** Emilie T. Théberge, Kate Durbano, Diane Demailly, Sophie Huby, Arezoo Mohajeri, Care4Rare Canada Consortium, Clara van Karnebeek, Gabriella A. Horvath, Karen Usdin, Anna Lehman, Laura Cif, Phillip A. Richmond

## Abstract

**Background:** Trinucleotide repeat expansions are an emerging class of genetic variants associated with several movement disorders. Unbiased genome-wide analyses can reveal novel genotype-phenotype associations and provide a diagnosis for patients and families.

**Objectives:** To identify the genetic cause of a severe progressive movement disorder phenotype in two affected brothers.

**Methods:** A family of two affected brothers and unaffected parents had extensive phenotyping and natural history followed since birth. Whole-genome and long-read sequencing methods were used to characterize genetic variants and methylation status. Results: We describe a CGG repeat expansion in the 5’-untranslated region of *DIP2B* in two affected male siblings presenting with a novel *DIP2B* phenotype including neurodevelopmental disability, dysmorphic traits, and a severe progressive movement disorder (prominent chorea, dystonia, and ataxia).

**Conclusions:** This is the first report of a severe progressive movement disorder phenotype attributed to a CGG repeat expansion in the *DIP2B* 5’-UTR.

## Introduction

A small fraction of short tandem repeats (STR) in the human genome have been associated with disease when one or both alleles exceed an STR-specific threshold in size^1^. Trinucleotide repeat disorders (TRDs) are a class of genetic diseases caused by STR expansions of 3 nucleotide repeat motifs, and the majority of currently known TRDs are diseases of the nervous system^2^. These TRDs can involve repeats located in both protein-coding regions such as seen in Huntington’s disease (OMIM #143100)^3^, and within noncoding regions, as seen in Friedreich’s ataxia (OMIM #229300), myotonic dystrophy 1 and 2 (OMIM #160900, #602668), and fragile X syndrome (FXS; OMIM #300624)^2^. Pathogenetic mechanisms for noncoding repeat expansions are incompletely understood and are actively being investigated across diseases and genomic loci.

A CGG trinucleotide repeat motif in the 5’ untranslated region (5’-UTR) of Disco-Interacting Protein 2 Homolog B (*DIP2B*; OMIM #611379) was first characterized by Winnepenninckx *et al*.^4^, who demonstrated how a large CGG repeat expansion caused gene silencing that resulted in FRA12A-type intellectual disability (OMIM #136630). Methylation of cytosine nucleotides in cytosine-guanine (CpG) dinucleotide pairs in CpG islands adjacent to promoters have been described to cause transcriptional gene silencing^5^. Since its initial association with FRA12A, a *DIP2B* 5’-UTR CGG expansion has been observed as the gene candidate in one individual with Lennox-Gastaut epilepsy^6^. There is not yet consensus on the number of repeats considered pathogenic for *DIP2B* expansion; it has been suggested that over 90 repeat copies may be of clinical relevance^7^, and ≤23 repeat copies more likely to be benign as observed from population-level genotyping in gnomAD^8^.

Herein, we report two male siblings with neurodevelopmental symptoms characterized by mild intellectual disability and a severe progressive motor disorder with chorea, dystonia, and ataxia. Short-read whole genome sequencing identified 79 and 96 copies of the CGG repeat motif in the 5’-UTR of *DIP2B* as the sole genetic candidate that could be responsible for this phenotype. Using Oxford Nanopore long-read whole genome sequencing (LRS) technology, we have now more accurately quantified the number of repeat motifs in affected and unaffected family members. We also showed that, unlike alleles with larger repeat numbers that are associated with intellectual disability, the expanded alleles are unmethylated.

## Methods

### Participants

Each family member provided informed consent for participation in research protocols approved by the University of British Columbia institutional ethics board (H12-00067 and H18-02912). Blood samples were collected for each family member in Montpellier, France, where the siblings were followed and underwent assessment for their movement disorder.

### Sequencing and bioinformatic analysis

The Canada’s Michael Smith Genome Sciences Center in Vancouver, Canada, generated the WGS and LRS results from peripheral whole blood samples. WGS was performed using Illumina HighSeqX (V2.5) on extracted DNA from the two brothers to investigate candidate genomic variants which could explain their shared phenotype: single nucleotide variants (SNVs), insertions and deletions (indels), and structural variants were prioritized after annotation for allele frequency in affected and general populations, and *in silico* predicted functional impact. Genes were prioritized according to previous association with neurological phenotypes, brain expression, and evidence of intolerance to variation. Repeat expansion genotyping was performed via ExpansionHunter^9^ and ExpansionHunter Denovo^10^ and visualized with REViewer^11^.

The Oxford Nanopore Promethion (P48) platform was used for LRS on all family members to fully genotype repeat expansions larger than the WGS read length, identify the parent of origin for the repeat expansion in each brother, and determine methylation status of *DIP2B*. LRS data were aligned to the GRCh38.p14 reference genome via minimap2^12^. Emerging tools in repeat expansion profiling were used: STRaglr^13^, TRGT^14^, vamos^15^, and Sniffles2^16^, and visualized via IGV and TRVZ^17^. Further details are described in the Supplemental Methods.

## Results

### Clinical course

Two brothers between 30-35 years old (probands II-1 and II-2) have been followed from their infancy for movement disorders of unknown etiology initially labeled as “cerebral palsy”, born following uneventful pregnancy and childbirth to healthy non-consanguineous parents of Sicilian ancestry (Figure 1A). Within their first year, both probands were observed to have developmental delay with central hypotonia. Fully autonomous standing and gait were never achieved for either proband. Between ages 1-3, both developed generalized chorea, with proband II-1 having the cranial region initially spared, unlike proband II-2. Initially, both probands had intelligible speech, behavioral milestones were achieved, and good quality social interactions were demonstrated. Both experienced mild cognitive impairments involving visuospatial processing and executive functions with altered inhibitory control, cognitive flexibility, working memory and attention span; intellectual disability was more prominent in proband II-2.

**Figure 1:**
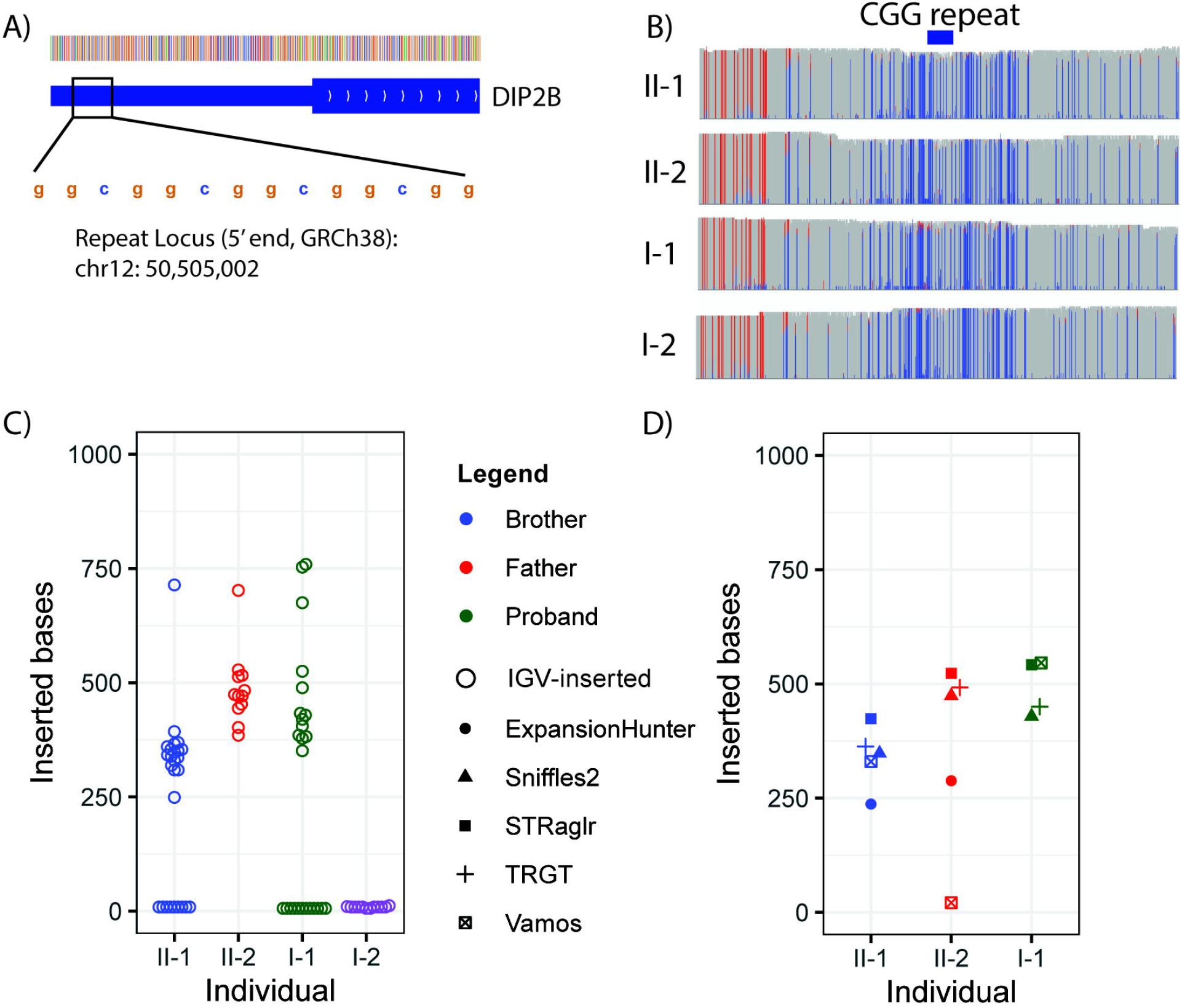
A) Two-generation pedigree. Filled black shapes indicated affected status, dot indicates unaffected carrier of *DIP2B* repeat expansion; B) T2-weighted axial brain MRI scans at comparable but not identical cut levels (columns: cerebellum, basal ganglia and lateral ventricles, and cortical levels) respectively of the proband and brother at two timepoints. The first series were done at late teenage years for both proband II-1 (I-III) and proband II-2 (IV-VI), without any obvious signal or structural alterations. Repeated scans 9 (proband II-1, VII-IX) and 10 (proband II-2, X-XII) years later show a non-specific pattern of mild, but significant for age, cerebellar and diffuse cortical atrophy without significant signal alterations. Observations indicated by circles include mild cerebellar atrophy for the two probands (yellow: panels I and VII and IV and X), non-specific pattern of diffuse cortical and hemispheric atrophy (red: panels II, VIII, III, IX, VI and XII), and a discrete but noticeable enlargement of the posterior horn of the lateral ventricles (blue: panel V, XI). The green arrow indicates the artifact of the deep brain stimulation (DBS) electrodes (panel XI). The two siblings underwent DBS surgery in an attempt to improve their movement disorder with little motor and no disability improvement.

Motor disorder phenomenology progressed through adulthood involving severe chorea, generalized dystonia and ataxia. In his mid-20s, proband II-1 had severe dysphonia and cerebellar dysarthria with variable amplitude and a high pitched-voice. Prehension and bimanual tasks have worsened in recent years, with milder impairment observed in proband II-2. Severe impairment of visual pursuit and saccadic eye movements were observed at the most recent clinical observations in both probands. Other shared features include severe hyperlordosis, genu recurvatum and mild facial dysmorphic traits becoming more apparent in adulthood.

Clinical expression varies between the two siblings as does disease progression. The siblings were not born with any obvious craniofacial differences as illustrated by the video sequences in infancy (Supplemental Videos), but more characteristic facial appearances developed over time, as reported in other neurodevelopmental disorders^18,19^. Serial brain MRIs taken ten years apart indicated progressive diffuse hemispheric and cerebellar atrophy (Figure 1B) without remarkable signal intensity alteration. Further details of the anthropometric measurements and clinical presentations along a timeline are provided in Supplemental Table 1.

### Genome-wide sequencing and methylation analysis

No candidate variants emerged from the initial WGS analysis of SNVs, indels, and structural variants. ExpansionHunter De Novo identified a GGC (CGG) repeat expansion in the 5’-UTR of *DIP2B* as the top ranked variant from an outlier test in both probands (Figure 2A). ExpansionHunter was used to genotype the *DIP2B* locus, identifying repeats of 79 and 96 copies (237 bp and 288 bp) for probands II-1 and II-2 (Figure 2D).

**Figure 2:**
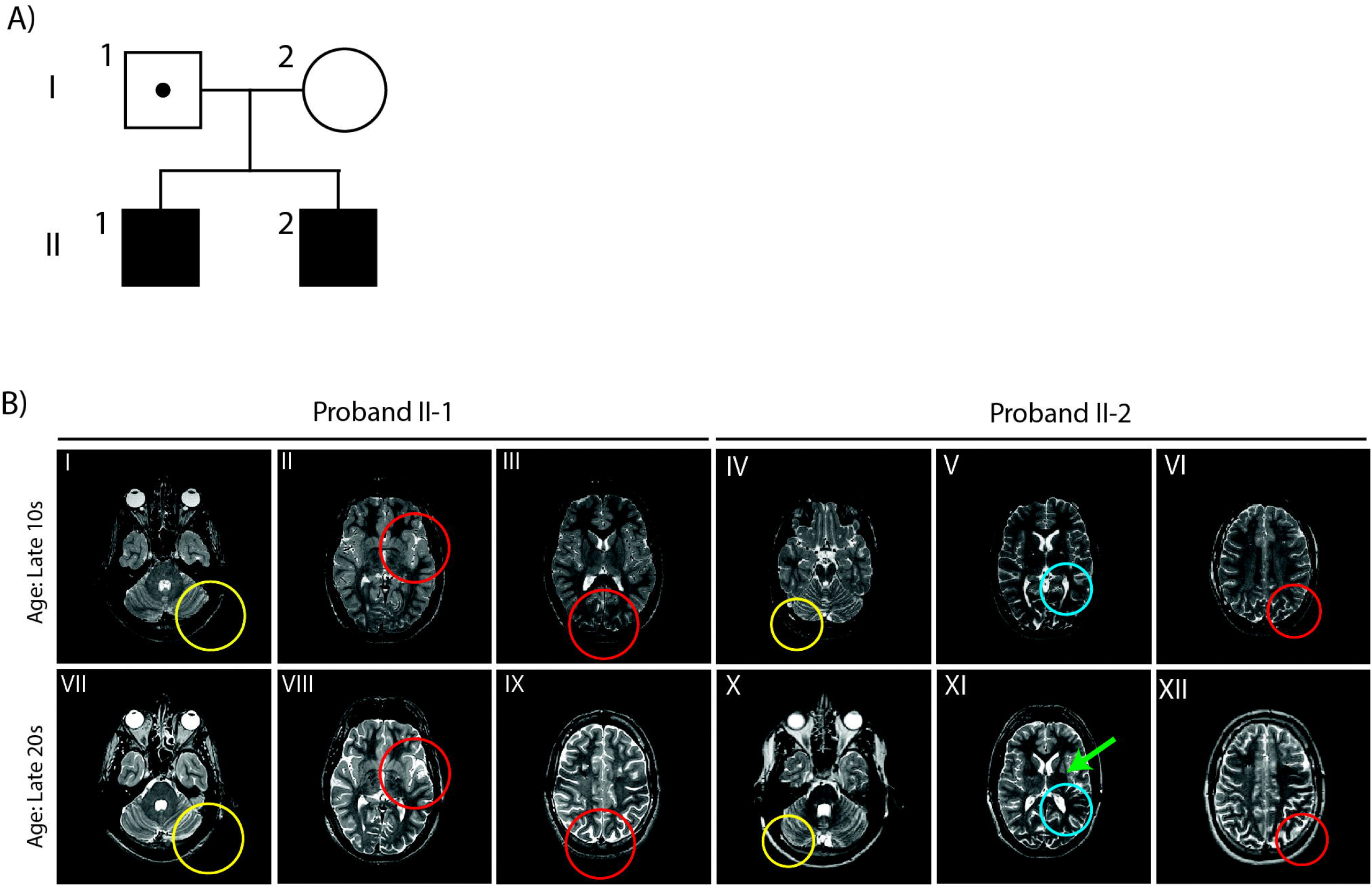
A) Visualization of the *DIP2B* genomic locus and GGC (CGG) repeat expansion in the 5’-untranslated region (UTR) region at chr12: 50,505,502 (1-based, GRCh38.p14); B) Histograms representing methylated (red) or unmethylated (blue) state of CpG dinucleotides across *DIP2B*; C) Number of inserted bases within aligned reads at the *DIP2B* locus for all four individuals; D) Variation in the number of *DIP2B* inserted bases called by ExpansionHunter (WGS), Sniffles2, STRaglr, TRGT, and Vamos (long read sequencing, LRS).

Visualization of the LRS data revealed that the repeat expansion with copy number greater than 100 CGG repeats was present in both probands as well as the father (Figure 2C, Supplemental Figure 1). Both probands were discordant for the maternal allele, ruling out the possibility of compound heterozygosity (Figure 2C). There is variability in the number of CGG repeats within each individual harboring the expansion (Figure 2C). There were no differences in CpG methylation frequency detected for either the repeat-flanking genomic regions (Figure 2B) or intra-repeat region (Supplemental Figure 2). Further, there were no interruptions detected in the CGG repeat tracks that may contribute to variable phenotype penetrance (e.g., AGG motif).

All of the tools applied to genotype the *DIP2B* CGG expansion were able to produce an expanded genotype across the probands and father, except for vamos, which failed to detect the expansion in proband II-2. The repeat count we identified is larger than those identified in recent population sequencing efforts using LRS^20^.

## Discussion

We have described a pair of male siblings with a severe, early onset prominent hyperkinetic and ataxic phenotype inherited from an apparently asymptomatic father. Our data implicate a CGG repeat expansion in the *DIP2B* 5’-UTR as the causative mutation. In murine animal models, *DIP2B* has been shown to be primarily expressed in the brain, with highest expression in the cerebral cortex, frontal and parietal lobes, and cerebellum^21^. DIP2B has been shown to regulate axonal guidance during early developmental stages, to play a role in regeneration in mature neurons, and also plays a role in hippocampal synaptic transmission morphogenesis^22^. Both probands present with mild intellectual disability associated with a severe progressive motor disorder as described above. Furthermore, serial brain MRI scans of the probands demonstrated diffuse cerebral and cerebellar atrophy. This study is the first to our knowledge to document progressive structural alterations in the human brain in two carriers of a *DIP2B* 5’-UTR CGG expansion.

Other CGG-expansion diseases with comparable sized CGG repeats at other loci are associated with symptoms similar to those seen in our probands, suggesting a possibly similar disease mechanism^23^. The observation of neurodegeneration in carriers of alleles with relatively small unmethylated CGG repeat tracts and intellectual disability in individuals with larger methylated expansions is reminiscent of what is seen in the Fragile X-related disorders, also caused by CGG repeat expansion, in this case in the 5’-UTR of the fragile X messenger ribonucleoprotein 1 gene (*FMR1*; OMIM #309550)^24^. FXS, a congenital developmental disorder characterized primarily by intellectual disability and other system impairments, is caused by the inheritance of an allele with >200 repeats. Like the *DIP2B* alleles associated with intellectual disability, these alleles become aberrantly methylated and are transcriptionally silent^25^. In contrast, alleles with 55-200 repeats are associated with an increased risk for Fragile X Tremor/Ataxia Syndrome (FXTAS; OMIM #300623), a disorder with variable penetrance that is associated with adult-onset cerebellar gait ataxia, intention tremor, and/or peripheral neuropathy^26^. FXTAS patients may also develop hypokinetic features such as parkinsonism and/or hyperkinetic symptoms. Late-onset intention tremor is the most frequently reported hyperkinetic movement disorder in FXTAS; however, chorea and dystonia have also been described^19^. FXTAS alleles, like the *DIP2B* alleles in our probands, are unmethylated^24^.

Although the *DIP2B* repeat has yet to be profiled at the population level using LRS, the gnomAD database^8^ reports 25 individuals out of 19,240 with ≥79 CGG-repeat copies at this *DIP2B* locus, a similar size observed in both probands from the short-read data. Larger LRS population studies will help to further profile this locus in its entirety and help establish an estimate of the penetrance for the expanded repeat genotype^20^.

With decreasing cost, improved accuracy, and increasing availability, LRS is emerging as a promising tool in the diagnosis of rare genetic diseases. In this case, LRS enabled the accurate profiling of the CGG repeat expansion in the *DIP2B* 5’-UTR, which we propose as a novel association with the shared movement disorder observed in the two affected probands. Recent work has demonstrated the etiologic roles of repeat expansions in patients afflicted by movement disorders, in particular ataxia^27,28^. Use of this technology within cases of undiagnosed movement disorder cohorts could both discover novel disease-associated loci and characterize pathogenic repeat expansions, ending the diagnostic odyssey for patients and families.

## Data Availability

Upon approval from the Care4Rare Canada Consortium, deidentified participant data with study accession numbers (Project 3058, samples BC200-203) may be available through access to Genomics4RD (genomics4rd.ca).

## Acknowledgements

The authors thank the probands and their family for their participation and consent to the publication. We are grateful to our colleagues in France for the follow-up of the patients, Dr. Solenne Correard for initial WGS analysis and in consenting translation aid, and colleagues at the Canada’s Michael Smith Genome Sciences Centre in Vancouver, Canada, for their sequencing work. This work was supported by grants awarded to C.V.K. and A.L. from the Canadian Institutes of Health Research, GenomeCanada and GenomeBC (University of British Columbia clinical research ethics board applications H12-00067 and H18-02912).

## Authors’ Roles

E.T.: Data collection coordination, writing, editing of final version of the manuscript.

K.D.: Data collection, data collection coordination, clinical patient interaction, validation of the final version of the manuscript.

D.D.: Data collection, clinical patient interaction, editing of final version of the manuscript.

S.H.: Data collection, clinical patient interaction, validation of final version of the manuscript. C.C.C.: Data management.

A.M.: Data management.

G.H.: Clinical expertise, data collection coordination, editing of final version of the manuscript. C.V.K.: Study oversight, editing of final version of the manuscript.

K.U.: Repeat expansion expertise, data interpretation, editing of final version of the manuscript. A.L.: Study oversight, clinical expertise, editing of final version of the manuscript.

L.C.: Clinical expertise, study oversight, clinical patient interaction, writing, editing of final version of the manuscript.

P.A.R.: Study oversight, data analysis, writing, editing of final version of the manuscript.

## Financial Disclosures of all authors

There are no financial disclosures or conflicts of interest reported by the authors related to the present manuscript. In the last 12 months:

E.T. receives employment income from the University of British Columbia (Vancouver, Canada).

K.D. receives employment income from the Hospital Center University De Montpellier (Montpellier, France).

D.D. receives employment income from the Beau Soleil Clinic (Montpellier, France)

S.H. receives employment income from the Hospital Center University De Montpellier (Montpellier, France).

A.M. receives employment income from the University of British Columbia (Vancouver, Canada)

G.H. receives employment income from the Provincial Health Services Authority (Vancouver, Canada).

C.V.K. receives employment income from the Amsterdam University Medical Centre (Amsterdam, Netherlands). Grants held include: European Joint Program, Rare Diseases 2021-

2025 EJP RD COFUND-EJP N° 825575; Horizon Health2023-2028: 101080249-2 SIMPATHIC; Metakids 2019-2024 United for Metabolic Diseases; and ZonMW 20120-2025 845008701 Transitional care unit.

K.U. receives employment income from the National Institutes of Health (Bethesda, USA).

A.L. receives employment income from the Provincial Health Services Authority (Vancouver, Canada). Research grants/clinical trial funding from Sanofi, Takeda, Idorsia; speaker fee from Takeda; on advisory board for Ultragenyx, Alexion, Sanofi and Amicus Therapeutics.

L.C. receives employment income from Department of Neurosurgery, University Hospital Montpellier, France. L.C. received consulting honoraria for educational activities from Boston Scientific and support from Canadian Dystonia Research Foundation outside of the submitted work.

P.A.R. receives employment income from BC Children’s Hospital Research Institute, Alama Health LLC, and Umoja Biopharma.

## Tables

No tables present in the main body of the text.

## References

1. Bahlo M, Bennett MF, Degorski P, Tankard RM, Delatycki MB, Lockhart PJ. Recent advances in the detection of repeat expansions with short-read next-generation sequencing. F1000Res. 2018;7(F1000 Faculty Rev):736.

2. Hannan AJ. Tandem repeats mediating genetic plasticity in health and disease. Nat Rev Genet. 2018;19: 286–298.

3. Tabrizi SJ, Estevez-Fraga C, van Roon-Mom WMC, et al. Potential disease-modifying therapies for Huntington’s disease: lessons learned and future opportunities. Lancet Neurol. 2022;21: 645–658.

4. Winnepenninckx B, Debacker K, Ramsay J, Smeets D, Smits A, FitzPatrick DR, Kooy, FR. CGG-repeat expansion in the DIP2B gene is associated with the fragile site FRA12A on chromosome 12q13.1. Am J Hum Genet. 2007;80: 221–231.

5. Cheung WA, Johnson AF, Rowell WJ, et al. Direct haplotype-resolved 5-base HiFi sequencing for genome-wide profiling of hypermethylation outliers in a rare disease cohort. Nat Commun. 2023;14: 3090.

6. Qaiser F, Sadoway T, Yin Y, et al. Genome sequencing identifies rare tandem repeat expansions and copy number variants in Lennox-Gastaut syndrome. Brain Commun. 2021;3: fcab207.

7. Mitina A, Khan M, Lesurf R, et al. Genome-wide enhancer-associated tandem repeats are expanded in cardiomyopathy. EBioMedicine. 2024;101: 105027.

8. Karczewski KJ, Francioli LC, Tiao G, et al. The mutational constraint spectrum quantified from variation in 141,456 humans. Nature. 2020 May;581(7809): 434–443.

9. Dolzhenko E, Deshpande V, Schlesinger F, et al. ExpansionHunter: a sequence-graph-based tool to analyze variation in short tandem repeat regions. Bioinformatics. 2019;35: 4754–4756.

10. Dolzhenko E, Bennett MF, Richmond PA, et al. ExpansionHunter Denovo: a computational method for locating known and novel repeat expansions in short-read sequencing data. Genome Biol. 2020;21: 102.

11. Dolzhenko E, Weisburd B, Ibañez K, et al. REViewer: haplotype-resolved visualization of read alignments in and around tandem repeats. Genome Med. 2022;14: 84.

12. Li H. Minimap2: pairwise alignment for nucleotide sequences. Bioinformatics. 2018;34: 3094–3100.

13. Chiu R, Rajan-Babu I-S, Friedman JM, Birol I. Straglr: discovering and genotyping tandem repeat expansions using whole genome long-read sequences. Genome Biol. 2021;22: 224.

14. Dolzhenko E, English A, Dashnow H, et al. Characterization and visualization of tandem repeats at genome scale. Nat Biotechnol. 2024. Advance online publication (2 Jan. 2024).

15. Ren J, Gu B, Chaisson MJP. vamos: variable-number tandem repeats annotation using efficient motif sets. Genome Biol. 2023;24: 175.

16. Smolka M, Paulin LF, Grochowski CM, et al. Detection of mosaic and population-level structural variants with Sniffles2. Nat Biotechnol. 2024. Advance online publication (2 Jan 2024)

17. Robinson JT, Thorvaldsdóttir H, Winckler W, Guttman M, Lander ES, Getz G, Mesirov JP. Integrative genomics viewer. Nat Biotechnol. 2011;29: 24–26.

18. Axelrod FB, Gold-von Simson G. Hereditary sensory and autonomic neuropathies: types II, III, and IV. Orphanet J Rare Dis. 2007;2: 39.

19. Lapostolle A, Delion T, Arnaud S, Manceau P, Degos B. FXTAS patient presenting as Huntington-like generalized chorea. Rev Neurol. 2021;177: 445–446.

20. Gustafson JA, Gibson SB, Damaraju N, Zalusky MP, Hoekzema K, Twesigomwe D, et al. Nanopore sequencing of 1000 Genomes Project samples to build a comprehensive catalog of human genetic variation. medRxiv. 2024. Preprint (7 Mar 2024).

21. Sah RK, Bahadar N, Bah FB, et al. Analysis of Dip2B Expression in Adult Mouse Tissues Using the LacZ Reporter Gene. Curr Issues Mol Biol. 2021;43: 529–542.

22. Xing Z-K, Zhang L-Q, Zhang Y, et al. DIP2B Interacts With α-Tubulin to Regulate Axon Outgrowth. Front Cell Neurosci. 2020;14: 29.

23. Ishiura H, Shibata S, Yoshimura J, et al. Noncoding CGG repeat expansions in neuronal intranuclear inclusion disease, oculopharyngodistal myopathy and an overlapping disease. Nat Genet. 2019;51: 1222–1232.

24. Nobile V, Pucci C, Chiurazzi P, Neri G, Tabolacci E. DNA Methylation, Mechanisms of Inactivation and Therapeutic Perspectives for Fragile X Syndrome. Biomolecules. 2021;11(2):296.

25. Bagni C, Tassone F, Neri G, Hagerman R. Fragile X syndrome: causes, diagnosis, mechanisms, and therapeutics. J Clin Invest. 2012;122: 4314–4322.

26. Leehey MA. Fragile X-associated tremor/ataxia syndrome: clinical phenotype, diagnosis, and treatment. J Investig Med. 2009;57: 830–836.

27. van Kuilenburg ABP, Tarailo-Graovac M, Richmond PA, et al. Glutaminase Deficiency Caused by Short Tandem Repeat Expansion in GLS. N Engl J Med. 2019;380: 1433–1441.

28. Chen Z, Tucci A, Cipriani V, et al. Functional genomics provide key insights to improve the diagnostic yield of hereditary ataxia. Brain. 2023;146: 2869–2884.

