## Supplemental Figures 1 and 2 for "*DIP2B* CGG repeat expansion in siblings with neurodevelopmental disability and progressive movement disorder"

### **Supplemental Figure 1**

IGV snapshot for the *DIP2B* locus showing the read pileups and inserted bases (purple) for each individual. Each row represents a read, with empty rows (flat grey bar) indicating reference points; variation in I-1 reflects heterozygosity of an expansion (351-759 insertion) with the shorter wild type allele (6-9 insertions); For individuals II-2 and I-2, absence of inserted bases indicates the reference allele; For proband II-1 the inserted bases are out of view and each read contains an expanded genotype or a 9bp genotype inherited from I-2; For the father I-1, each read contains the expanded allele or a 6bp allele.

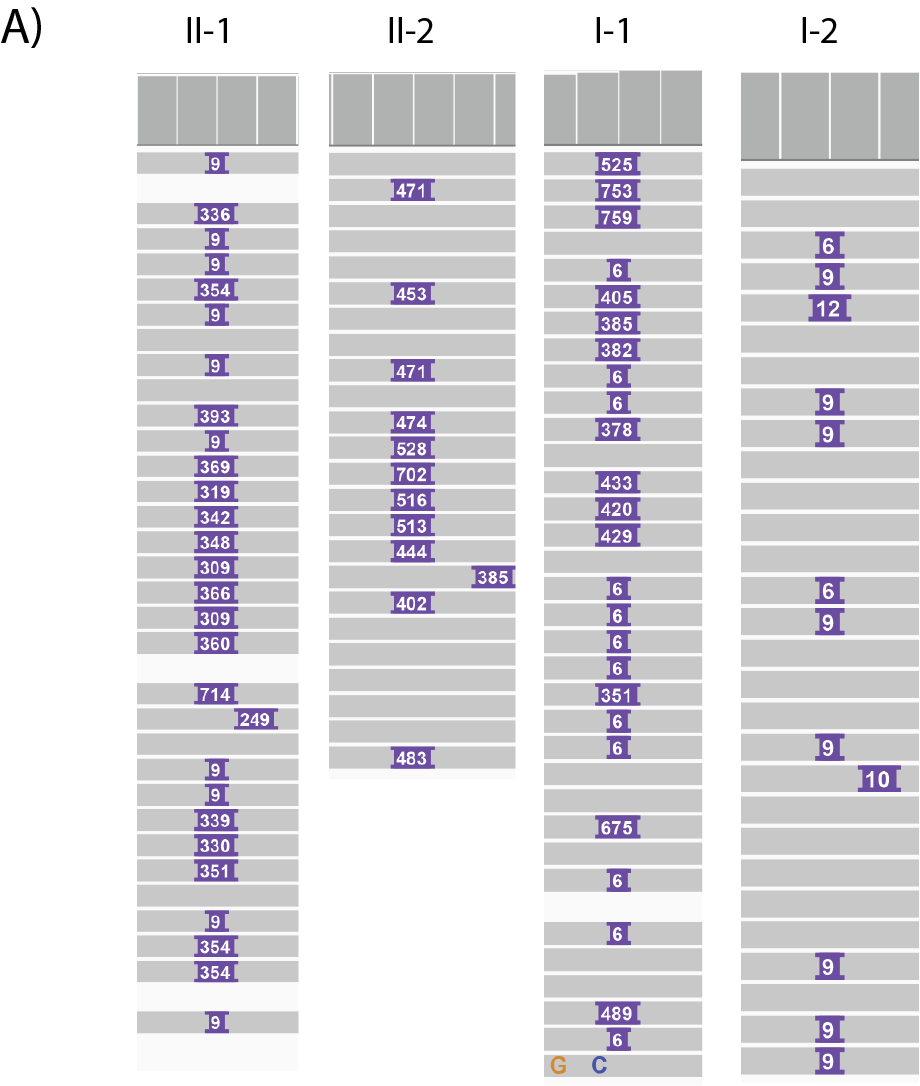

### **Supplemental Figure 2**

Waterfall plots showing the expanded and non-expanded reads across all individuals. Methylated CpG shown in red, and unmethylated CpG shown in blue.

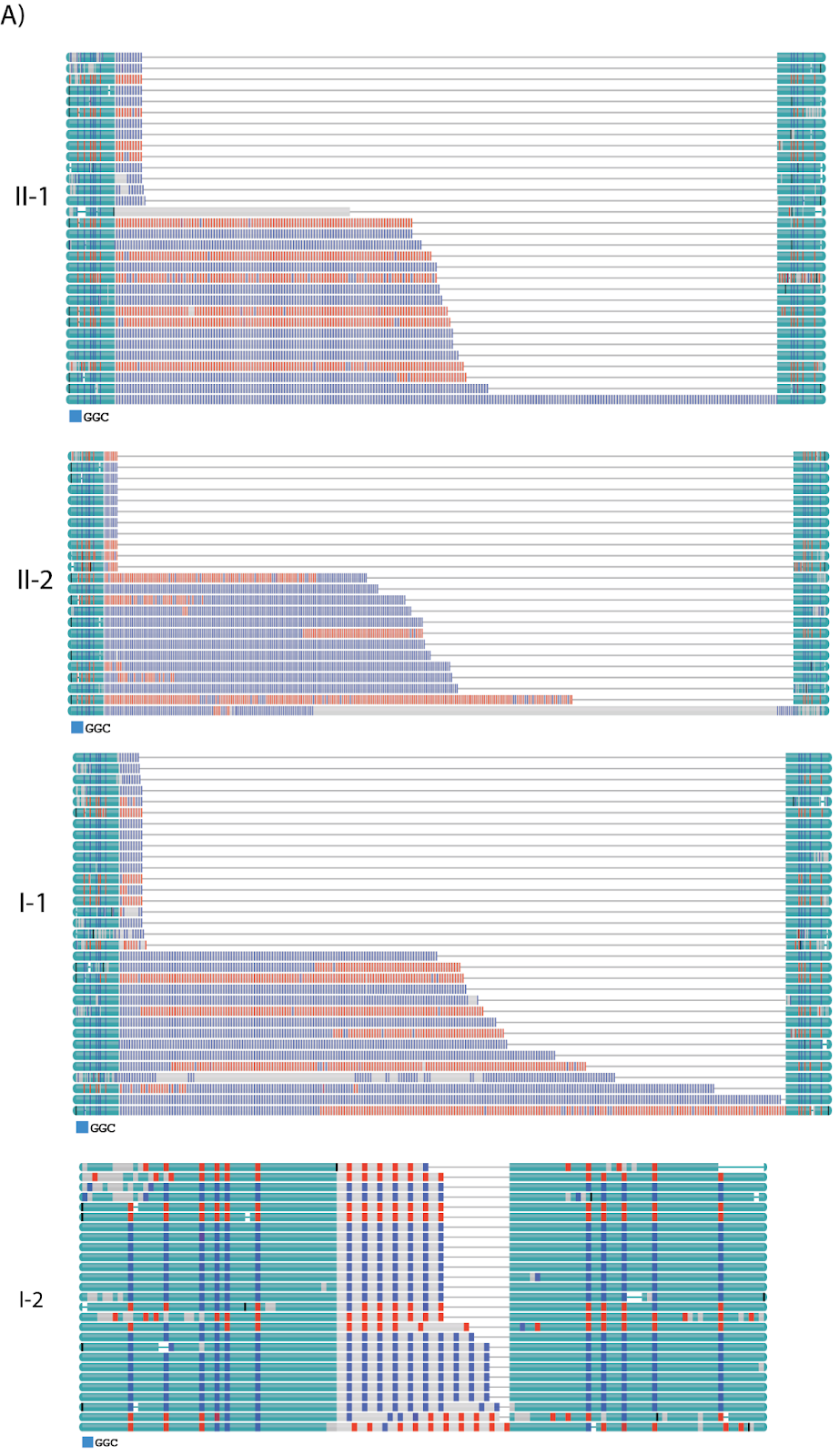
