## Supplemental Methods for "*DIP2B* CGG repeat expansion in siblings with neurodevelopmental disability and progressive movement disorder"

### **Supplemental Methods: Bioinformatic Analyses**

Short-read WGS was processed for both proband II-1 and II-2 as previously described^1^. Briefly, reads were aligned against the GRCh38.p14 reference genome using bwa mem (v0.7.17)^2^, converted using samtools (v1.10) to a binary format^3^, duplicate marked with picard (v2.26.3)^4^, SNV and indel variant calling with DeepVariant (v1.4.0)^5^, converted using BCFtools (v1.10)^6^, annotated with slivar (v0.2.1)^7^, vcfanno (v0.3.2)^8^ and VEP (v101.0)^9^, converted to a database using VCF2DB^10^, and queried using GEMINI (v0.30.2)^11^. SNVs and indels related to the ataxia phenotype were queried from HPO^12^ and MeSHOP^13^ associations from annotated variant tables as well as via the Exomiser tool (v12.1.0)^14^. Repeat expansions were profiled using ExpansionHunter De Novo (v0.9.0)^15^ and ExpansionHunter (v5.0.0)^16^, and visualized with REViewer (v0.2.7)^17^. For EHdn, a control cohort database derived from the 1000 Genomes project was used.

Long-read WGS was basecalled using guppy (v6.3.8, config dna_r10.4.1_e8.2_400bps_modbases_5mc_cg_sup_prom)^18^, aligned against the GRCh38.p13 no-alt reference genome using minimap2 (v2.22)^19^, converted file formats using samtools (v1.11)^3^, and visualized with IGV (v2.16.0)^20^. Repeat expansions were called using Sniffles2 (v2.2)^21^, vamos (v1.3.6)^22^, TRGT (v0.8.0)^23^, and STRaglr (v1.4.1)^24^. All tools were used with default settings, except for vamos which did not have a repeat expansion for the DIP2B locus in its catalog. A custom configuration for vamos was used which defined the repeat expansion in the same manner as TRGT giving the coordinates for the motif and the GGC motif (chr12:50505001-50505022, GGC). Repeat expansions were viewed in waterfall plots using the TRVZ tool (v0.8.0)^25^ and IGV.

**References**

1. van Kuilenburg ABP, Tarailo-Graovac M, Richmond PA, et al. Glutaminase Deficiency Caused by Short Tandem Repeat Expansion in GLS. N Engl J Med. 2019;380: 1433–1441.
2. Li, H. Aligning sequence reads, clone sequences and assembly contigs with BWA-MEM. arXiv:1303.3997
3. Li H, Handsaker B, Wysoker A, et al. The Sequence Alignment/Map format and SAMtools. Bioinformatics. 2009;25: 2078–2079.
4. Picard Toolkit. 2019, Broad Institute, Github Repository. <http://broadinstitute.github.io/picard>
5. Poplin R, Chang P-C, Alexander D, et al. A universal SNP and small-indel variant caller using deep neural networks. Nat Biotechnol. 2018;36: 983–987.
6. Danecek P, Bonfield JK, Liddle J*,* et al*.* Twelve years of SAMtools and BCFtools. Gigascience. 2021;10(2):giab008.
7. Garrison E, Kronenberg ZN, Dawson ET, Pedersen BS, Prins P. A spectrum of 788 free software tools for processing the VCF variant call format: vcflib, bio-vcf, cyvcf2, 789 hts-nim and slivar. PLoS Comput. Biol. 2022;18: e1009123.
8. Pedersen BS, Layer RM, Quinlan AR. Vcfanno: fast, flexible annotation of genetic variants. Genome Biol. 2016;17: 118.
9. McLaren W, Gil L, Hunt SE, et al. The Ensembl Variant Effect Predictor. Genome Biol. 2016;17: 122.
10. Pedersen, BS. Vcf2db. 2018, Github Repository. <https://github.com/quinlan-lab/vcf2db>
11. Paila U, Chapman BA, Kirchner R, Quinlan AR. GEMINI: integrative exploration of genetic variation and genome annotations. PLoS Comput Biol. 2013;9: e1003153.
12. Köhler S, Gargano M, Matentzoglu N, et al. The Human Phenotype Ontology in 2021. Nucleic Acids Res. 2021;49: D1207–D1217.
13. Cheung WA, Ouellette BF, Wasserman WW. Inferring novel gene-disease associations using Medical Subject Heading Over-representation Profiles. Genome Med. 2012; Sep 28;4(9):75.
14. Smedley D, Jacobsen JOB, Jäger M, et al. Next-generation diagnostics and disease-gene discovery with the Exomiser. Nat Protoc. 2015;10: 2004–2015.
15. Dolzhenko E, Bennett MF, Richmond PA, et al. ExpansionHunter Denovo: a computational method for locating known and novel repeat expansions in short-read sequencing data. Genome Biol. 2020;21: 102.
16. Dolzhenko E, Deshpande V, Schlesinger F, et al. ExpansionHunter: a sequence-graph-based tool to analyze variation in short tandem repeat regions. Bioinformatics. 2019;35: 4754–4756.
17. Dolzhenko E, Weisburd B, Ibañez K, et al. REViewer: haplotype-resolved visualization of read alignments in and around tandem repeats. Genome Med. 2022;14: 84.
18. Wick RR, Judd LM, Holt KE. Performance of neural network basecalling tools for Oxford Nanopore sequencing. Genome Biol. 2019 Jun 24;20(1):129.
19. Li H. Minimap2: pairwise alignment for nucleotide sequences. Bioinformatics. 2018;34: 3094–3100.
20. Robinson JT, Thorvaldsdóttir H, Winckler W, Guttman M, Lander ES, Getz G, Mesirov JP. Integrative genomics viewer. Nat Biotechnol. 2011;29: 24–26.
21. Smolka M, Paulin LF, Grochowski CM, et al. Detection of mosaic and population-level structural variants with Sniffles2. Nat Biotechnol. 2024 Jan 2. Epub ahead of print.
22. Ren J, Gu B, Chaisson MJP. vamos: variable-number tandem repeats annotation using efficient motif sets. Genome Biol. 2023;24: 175.
23. Dolzhenko E, English A, Dashnow H, et al. Characterization and visualization of tandem repeats at genome scale. Nat Biotechnol. 2024. 2024 Jan 2. Epub ahead of print.
24. Chiu R, Rajan-Babu I-S, Friedman JM, Birol I. Straglr: discovering and genotyping tandem repeat expansions using whole genome long-read sequences. Genome Biol. 2021;22: 224.
