## Supplemental Table 1 for "*DIP2B* CGG repeat expansion in siblings with neurodevelopmental disability and progressive movement disorder"

### **Supplemental Table 1: Phenotypic Progression**

Phenotypic description for Proband II-1 and Proband II-2 from pregnancy and birth through adulthood.

| **Age (in years)** | **Proband II-1** | **Proband II-2** |
| --- | --- | --- |
| **Pregnancy and birth** | Uneventful pregnancy and birth; Apgar score 10/10  Weight~3480 g; Height~50 cm; Head circumference, 33 cm | Uneventful pregnancy and birth; Apgar score 10/10  Weight~3150 g; Height~49 cm; Head circumference, 33 cm |
| **0-1** | *Delay in developmental milestones* observed but not reported. | *Delay in developmental milestones* observed and reported by the age of 6 months. |
| **2-3** | **Milestones**: Able to turn in bed; unsteady sitting-up. *Standing and gait not achieved (few non autonomous steps).* Behavior adapted for age.  **Movement disorder**: *Abnormal lower limb posturing.*  **Milestones**: Grasps bilaterally and holds an object; Says words to label a person or object; Understands and executes simple orders, mimicking eating, and drinking; answers to simple questions.  **Movement disorders**: *Generalized chorea but cranial region spared.* | **Milestones:** Autonomous turning in the bed, crawling, steady autonomous sitting up. Bilateral grasping and holding objects achieved. *Autonomous standing and gait not achieved (walks with support)*. Behavior fully adapted to age and good quality social interactions.  **Movement disorders**: *Generalized chorea.* |
| **4-10** | **Movement disorders:** Prominent generalized chorea, involvement of cranial segment; Can stand-up with support and walk with a rollator/walker.  Persistence of generalized chorea. Speech intelligible. | Generalized chorea increased in intensity. *Mild dystonic component involving the cranial segment.* |
| **11-20** | Motor regression, loss of capacity for sitting up, bilateral support needed for standing. **Movement disorders**: *Dystonic postures at four limbs.* Hyperlordosis and genu recurvatum. Generalized chorea. *Cerebellar dysarthria with amplitude variation and high-pitched voice.* | Generalized chorea increasing with posture maintain and action. Dystonic postures involving the limbs distally. Limb ataxia. Cerebellar dysarthria with amplitude variation and high-pitched voice. |
| **>21** | Grasping difficult. Severe combined movement disorder with generalized dystonia and chorea* involving the cranial segment and *laryngeal dysphonia*, cerebellar dysarthria and chorea. *Altered pursuit and altered saccades (onset at 25y).* Brisk tendon reflexes at 4 limbs.  *Mild improvement of chorea under Tetrabenazine. | Chorea at rest is absent but elicited by posture maintain and by action. Generalized right side prominent dystonia involving the cranial segment. Limb ataxia and dysmetria. *Altered pursuit and saccadic movements*. Hyperlordosis and genu recurvatum. Standing-up and gait are non-autonomous. |
